# Use of nudge techniques in General practice (GP) to promote HPV vaccination among parents of boys aged 11-14 years: effectiveness and acceptability

**DOI:** 10.1101/2022.09.19.22280024

**Authors:** Adriaan Barbaroux, Morgane Chanzy

## Abstract

**Background:** A nudge is any procedure that modifies and/or influences the architecture of a choice, without using persuasion or financial incentives. It is commonly referred to as a “psychological nudge”. Nudges are effective in promoting public health issues such as HPV vaccination. Several systematic reviews of the literature place nudges among the most promising interventions for increasing vaccination coverage.

**Objective:** The objective of this study is to evaluate the effectiveness and acceptability of a nudge towards HPV vaccination based on the proposal of a consultation appointment to the parents of boys aged 11 to 14 years in the patient base of two general practitioners. The secondary objective is to evaluate the role of the feeling of control in this acceptability.

**Methods:** Participants were divided into two parallel experimental arms: a nudge group and a non-nudge group. The procedure used was a simple randomization of the parents of boys aged 11-14 years into two groups in the general practitioner’s practice. The study was conducted in two phases and took the form of a proposed appointment at the private practice of the participants in the nudged group. A questionnaire was administered to the participants in both groups one month after these appointments, asking them about their vaccination status regarding HPV vaccination, their acceptability of the nudge and their feeling of control in this procedure.

**Results:** The analysis was carried out on 32 participants in each of the two groups. The nudge was significantly effective in terms of vaccination coverage. Similar acceptability was found in both groups. The absence of a feeling of control was noted.

**Conclusion:** This study shows that the prevalence of a vaccination can be significantly modified by a nudge. This study did not show a significant difference in the acceptability of the nudge in the two groups. However, the acceptability of the nudge is significant in both groups. The literature shows good external consistency across different populations in France. This could mean that the French population is predominantly pro-nudge. Thus, a nudge deemed acceptable by the participants is not necessarily ethical, and may insidiously implant an idea. The ethical approach to nudges before their implementation is therefore essential.

## Introduction

### Background

HPV vaccination coverage for boys and girls aged 11-14 years remains well below target despite numerous recommendations, promotional campaigns and data on vaccine efficacy and safety. In 2020, the complete vaccination coverage of girls aged 16 was 32.7%, whereas a minimum vaccination rate of 60% of this population is needed to significantly reduce the occurrence of cancer in the overall population (1). Boys coverage is even lower, raising the question of how to encourage vaccination among the parents of young boys and among adolescents.

Nudging is an effective and inexpensive method of increasing people’s adherence to a health behaviour without persuasion or financial incentive. It was proposed by Thaler and Sunstein in 2008 and defined as a *gentle method of inspiring the right* decision (2). Nudge refers to any modification of the architecture of choice that aims to change people’s behaviour in a predictable way without disallowing them any options or changing their motivations (3). Nudging is considered to be one of the most efficient techniques for changing simple behaviours (4).

Here, the nudge has been carried out as an “opt-out”. According to a study in the UK, “an opt-out system” is likely to increase “the rate of consent for organ donation after death”. There are two broad categories of legal framework for consent to organ donation after death: “opt-in, where the explicit consent of the patient or his or her relatives is required, and opt-out, where consent is presumed in the absence of a statement to the contrary made by the patient during his or her lifetime. Since the introduction of an opt-out in 2020 in England, there has been a significant increase in organ donation. In this study, contacting a participant by telephone without their prior consent constitutes the opt-out. However, the participant must be informed of his or her right to withdraw from the study following this contact. These are the most effective nudges in the literature (5).

Choice architecture describes how decisions are influenced by the presentation of different possibilities. People can be steered in a certain directions by the choice architecture without restricting their individual freedom of choice. For example: placing healthy foods in a school cafeteria at eye level and placing less healthy foods in a less accessible place is a simple example of nudging. Individuals are still able to choose what they like, but by the way the food is organized, they are encouraged to eat healthier food (6). The nudge is unconscious in this situation.

This process, which has been studied for years in social psychology, has received renewed attention since Richard Thaler was awarded the Nobel Prize in Economics in 2017 for his work on the application of nudging to economics. The efficiency of nudging is recognized for several applications such as the promotion of organ donation, healthy eating or vaccination (7). Several studies have shown the effectiveness of nudging in increasing vaccination coverage (8) and some authors describe nudges as one of the most effective interventions (9).

Several factors influence the acceptability of nudges (22). A questionnaire study of a thousand participants in Sweden and the United States showed that attitudes vary according to the type of nudge: those with a direct benefit for the individual “pro-self nudge” were more accepted than those without a direct benefit for the individual “pro-social nudge” (8). Acceptability also varied according to the type of cognitive functioning of the individuals: a worldview focused on the benefit of society and rational reasoning were associated with better acceptability. Finally, Swedes found nudges more acceptable than North Americans (10).

Including educational elements about HPV infection in addition to information about access to vaccination services increases the acceptability of vaccination. Risk knowledge can enhance perceptions of self-efficacy and improve vaccination coverage in the population (11).

For this reason, the nudge studied here consisted in offering an HPV vaccination information consultation appointment and not a vaccination consultation per se.

Boys are less exposed to information about vaccination even though they are also at risk of HPV-related cancer. Improving parents’ knowledge via a dedicated consultation could reduce this inequality and contribute to the fight against HPV-related cancers. If the effectiveness of nudges is demonstrated, vaccination coverage of adolescents in France could be increased globally by campaigns based on nudges, and boys who would otherwise not have been vaccinated will be protected and will be able to protect their future sexual partner(s).

### Theoretical framework and hypothesis

The objective of this study was twofold: to assess the effectiveness and acceptability of an opt-out nudge aimed at increasing vaccinations of 11-14 year old boys in the GP population. Parents of boys aged 11-14 years in the general practice population were surveyed for the study.

The main hypothesis was that the effectiveness of the nudge would be such that the vaccination rate would be higher in the group exposed to the nudge than in the unexposed group (hypothesis 1a) and that the acceptability of the nudge would be better in the subjects exposed to the nudge (hypothesis 1b). In addition, in this study, particular attention was paid to the feeling of control of participants in both groups (nudged and non-nudged) regarding the telephone call, the appointment and the possible vaccination. It was assumed that both groups did not feel a loss of control in the choice of whether or not to make the appointment and whether or not to be vaccinated (hypothesis 2).

## Materials and Methods

### Setting, participants, design and procedure

This unicentric, two-arm parallel trial was designed to control for the effectiveness and acceptability of nudges. A simple and equal randomization (1:1 for both groups) was used. The participants were parents of boys aged 11 to 14 years and were divided into two parallel experimental arms: a nudge group and a no-nudge group. Recruitment was carried out in two general practices in southern Corsica. Participants were contacted by telephone via the GPs’ patient database. The procedure used was a simple randomisation of the parents of boys aged between 11 and 14 years in the patient base of one or more general practitioners into two groups by random number table. The study was conducted in two phases.

Phase 1 consisted of contacting the participants of the nudged group by telephone. They were offered a consultation and information appointment at a predefined date and time in order to remove the barriers to making an appointment. The consultation took place at their GP’s surgery. A prescription including the Gardasil 9 vaccine and an information document summarising the points discussed during the appointment were given to the participants (12). Only participants in the nudged group were contacted at this stage. They were explicitly informed that they were nudged. They were given the opportunity to postpone or cancel their appointment. Oral consent (no objection) was then taken at this point. A confirmation text message including the name of the study, the date and time of the information appointment was sent to them following the telephone call. A special note was included in the text message so that participants who wanted to leave the study could do so simply by texting “STOP” in response to the appointment confirmation message.

Phase 2 consisted of a telephone call to the nudged group one month after their exposure to the nudge. A questionnaire was administered during the telephone call. The questionnaire assessed their HPV vaccination status following exposure to the nudge. The non-nudged group was contacted for the first time during the second phase of the study. They were not offered an appointment. They answered a questionnaire similar to that of the nudged group, asking about their vaccination status with regard to HPV and their acceptability of the nudge performed on the nudged group. Their sense of control was also assessed using a verbal scale of “a little, a lot, not at all”.

The selection criteria for participants were parents of boys aged 11 to 14 years found in a general practitioner’s patient file. The required number of subjects was calculated using the “BiostaTGV” website. In order to obtain a statistical power of 90% with a tolerated alpha risk of 5%, 46 residents per group were required for the efficacy study, assuming a vaccination rate of 60% in the incentive group versus 30% in the non-incentive group. The criteria for non-inclusion of participants were participants who could not be contacted by telephone. The exclusion criteria were the participants from the study who refused to enroll.

The data was entered anonymously into an Excel spreadsheet. A second Excel spreadsheet containing the identity and telephone number of each subject was created. A matching spreadsheet between each subject and the random numbers identifying the corresponding data set was created to allow deletion of data from participants wishing to leave the study. The spreadsheets were stored on encrypted keys using Veracrypt® encryption software. The data were collected through the use of patient records on GPs’ secure software: “Hellodoc” containing information not available to the general public.

There is an arm’s length relationship between the participants and the study investigators. The research did not include people who are in a subordinate relationship with the study’s investigators. The participants were informed at the beginning of the study that they were nudged. The objectives of the study and its implications were detailed to the participants from the outset. The various forms are available in the appendix.

### Data analysis

The focus of the study was on the effectiveness and acceptability of the nudge as well as on perceived control. The independent variables were exposure to the nudge and the vaccination status of the nudged group for the acceptability study.

The effectiveness of the nudge on vaccination rates was assessed by χ2 tests. The consistency of the nudge acceptability scales was tested using Crohnbach’s alpha. All statistical analyses were performed using JASP software.

### Regulatory and ethical aspects

The study was carried out in accordance with the French regulations in force at the time of data collection. As an interventional study in the human and social sciences applied to the field of health, the study was classified outside the scope of the Jardé law by the Comité de protection des Personnes (CPP) of Saint Etienne (ID RCB number: 2021-A01140-41 10.05.2021). The project was then submitted to the Collège National des Généralistes Enseignants (CNGE) as well as to the Comité Éthique pour les Recherches Non Interventionnelles (CERNI) of the Université Côte d’Azur (Number AVIS n° 2022-018) which issued a favorable opinion. This work was the subject of a declaration of compliance with the reference methodology 004 to the CNIL n°2221990v0.

## Results

### Descriptive statistics

Phase 1 took place between 25 April and 19 May 2022. Phase 2 took place between 25 April and 15 June 2022. Of the 132 patients recruited from the GP database, 67 were excluded because they could not be contacted by telephone. No telephone numbers were found in the files for 38 of these patients, nine numbers were no longer assigned, 20 patients did not answer the call despite three attempts on three successive days. A flow chart is available in Figure 1. All participants included in Phase 1 were able to be contacted again in Phase 2 of the study. One participant in the non-nudge group asked to leave the study. In the end, 64 participants were surveyed: 32 participants in the nudge group and 32 in the non-nudge group (fig. 1). The boys in the participants were all aged between 11 and 14 years: 16 boys aged 11 years (25%), 17 boys aged 12 years (26.5%), 17 boys aged 13 years (26.6%) and 14 boys aged 14 years (21.9%).

**Figure 1.**
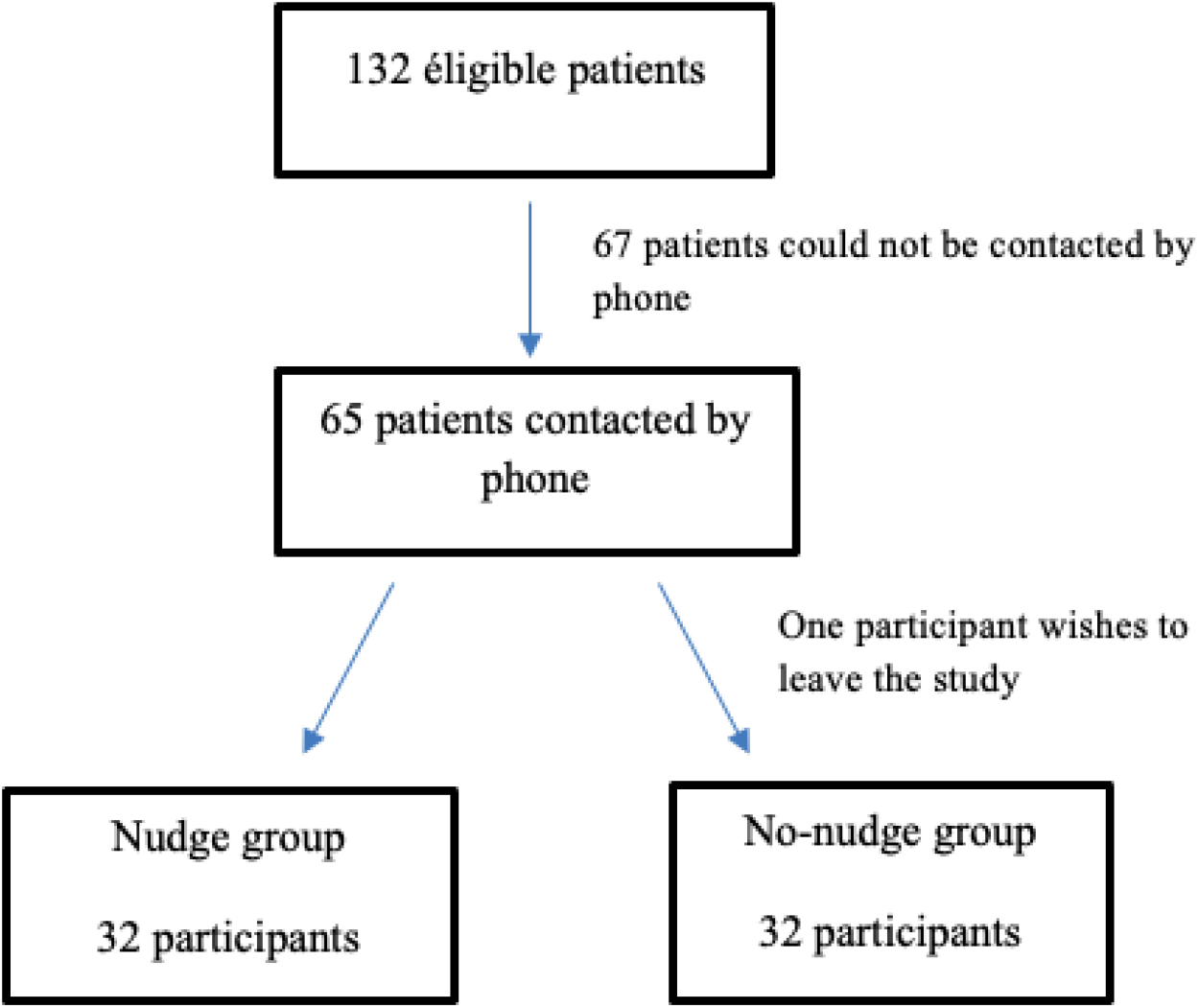
Flow chart.

Before the call, 87.5% of participants knew that vaccination was recommended for girls and 34.4% knew that vaccination was recommended for boys. Of the 64 participants, 62 considered their child’s vaccinations to be up to date and two did not know.

### Nudge Effectiveness

Of the nudged group, 17 (53%) said their child had been vaccinated against HPV, compared to 3 (9.4%) in the non-nudged group. This difference was statistically significant (*p* < 0.005). Among the 42 parents of unvaccinated children, 26 (61.9%) stated that they intended to have their child vaccinated this year. This proportion was significantly higher in the nudged group (94%) than in the non-nudged group (53%), *p* < 0.01.

**Table 1.** Effectiveness of nudge in 64 patients followed in general practice, divided into two groups: nudge and no-nudge. Vaccination status of the nudged group in phase 1 compared to phase 2 (post exposure to the nudge) vs vaccination status of participants in the non-nudged group contacted in phase 2 of the study. South Corsica, France 2022 (N = 64).

|  | Nudge group<br>(n = 32) | No-nudge group<br>(n = 32) |
| --- | --- | --- |
| Vaccinated before phase 1, N<br>(percentage) | 2 (6,2 %) | Data not collected |
| Vaccinated in phase 2, *i.e.* after<br>exposure to nudge for the nudged<br>group, N (percentage) | 15 (50 %) | 3 (9,4 %) |
| Reported intention to be vaccinated<br>in the year, N (percentage) | 15 (100 %) | 10 (53 %) |
| Total (N=64) | 32 | 13 |

### Acceptability of nudge

Crohnbach’s alpha was 0.82 for all acceptability subscales, which corresponds to good consistency. Both groups rated the nudge as very acceptable with a mean of 4.7 on a 5-point Likert scale ranging from 1 (not at all) to 5 (Totally acceptable) in the non-nudged group and 4.9 in the NN group. This difference was not significant (*p* = 0.15). Acceptability, on the other hand, was significantly influenced by adherence to current recommendations (*p* < 0.001, *F* = 5.5, corresponding to a high effect size (13).

The questionnaire asked whether participants would like to be nudged in the future. A quarter of the participants did not know how to answer the question “In the future, would you like to be offered a vaccination appointment straight away? One person in the nudged group answered no and three people in the non-nudged group answered no. This difference was not statistically significant (*p* = 0.16) but the non-nudged group had three times as many ‘don’t know’ responses as the nudged group (*p* < 0.05).

**Table 2.**
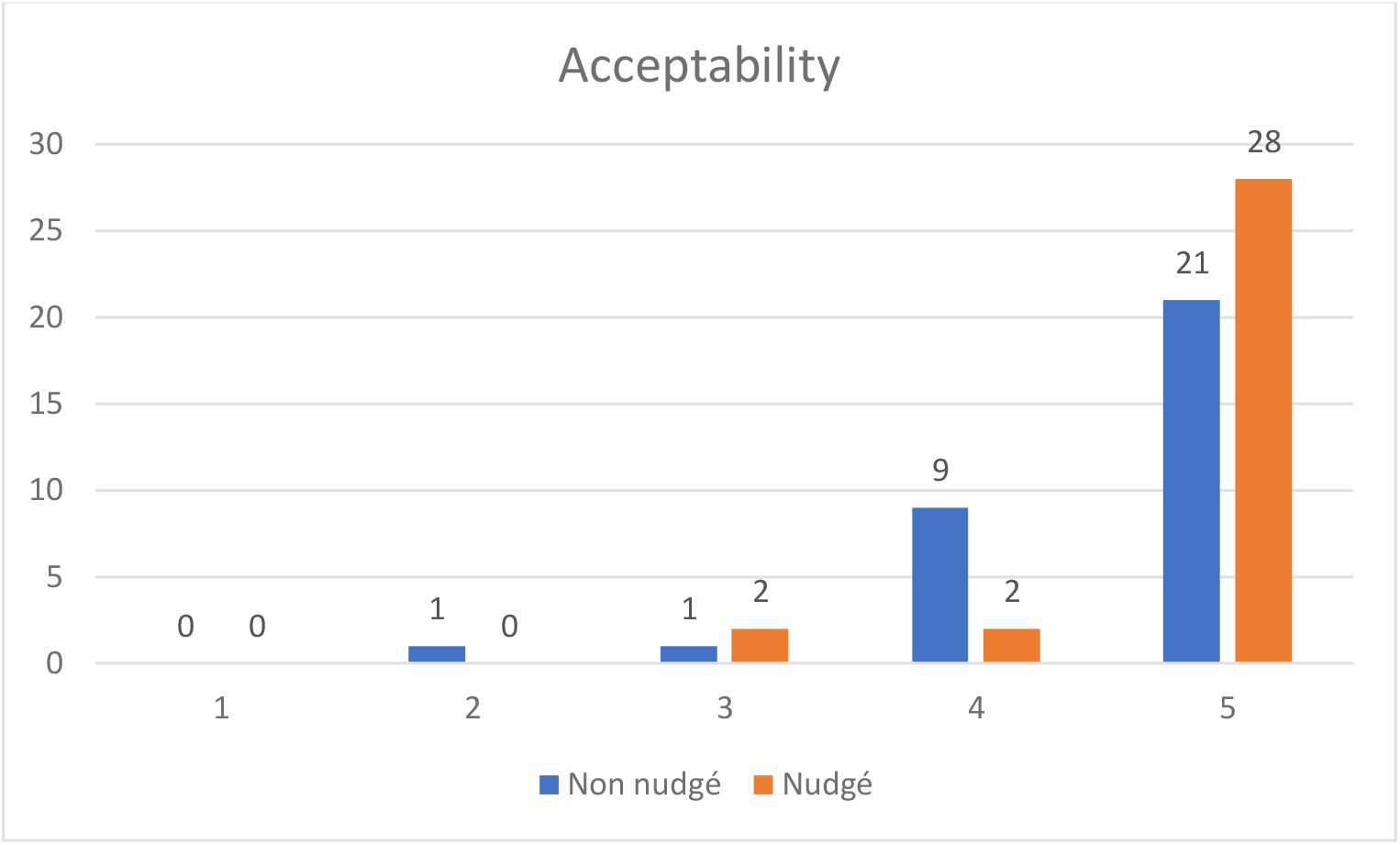
Acceptability of nudge in 64 patients followed in general practice, divided into two groups: nudge and no-nudge. On the ordinate, the number of participants. On the x-axis, the scale corresponding to 1: Not at all acceptable, 5: Totally acceptable. South Corsica, France 2022.

**Table 3.** Acceptability of nudge in 64 patients followed in general practice, divided into two groups: nudge and no-nudge. Average results per group. South Corsica, France 2022. (N=64)

| SCALE | ACCEPTABILITY |  |  |
| --- | --- | --- | --- |
|  | Nudge<br>(n=32) | No nudge<br>(n=32) | TOTAL<br>(N=64) |
| 1 Not at all acceptable | 0,0% | 0,0% | 0,0% |
| 2 Not acceptable | 0,0% | 3,1% | 1,6% |
| 3 Moderately acceptable | 6,3% | 3,1% | 4,7% |
| 4 Fairly acceptable | 6,3% | 28,1% | 17,2% |
| 5 *Totally acceptable* | 87,5% | 65,7% | 76,6% |

### Participants’ perception of control

All the participants said that this procedure did not deprive them of control, neither over the choice of appointment, nor over whether to vaccinate their son or not.

### Post-hoc analysis (additional analyses)

Nudged individuals were more likely to advise others to vaccinate themselves or their children (M = 3.6 Vs 4.2; *p* < 0.05). However, the degree of adherence to vaccine recommendations did not differ in these two groups (M = 3.8 Vs 4.2; *p* = 0.15). In other words, simply being exposed to a nudge promoting HPV vaccination appears to increase adherence to HPV vaccination, without significantly influencing adherence to vaccination recommendations in general.

## Discussion

In line with the main hypothesis (h1a), the results of this study showed that the nudge was significantly effective in increasing the prevalence of HPV vaccination. It was not an unconscious inducement to vaccinate because participants knew they were being nudged.

Contrary to our main hypothesis (h1b), there is no significant difference in the acceptability of nudge in the two groups. However, it is important to note that the acceptability of nudge is significant in both groups. However, other studies on nudge (14,15) have shown that the social acceptability of a nudge is artificially increased by the prior exposure of the population to a nudge. This was the assumption of the cognitive dissonance study(16). However, here the participants were informed that they were being nudged, which could lead to the assumption that they did not experience cognitive dissonance and therefore that the nudges were really accepted by the participants. A number of studies have described the social acceptability of nudges as generally correct: nudges deemed acceptable by 40 to 87% of participants (10).

The literature shows good external consistency across several different populations in France. This could mean that the French population is predominantly pro-nudge.

In line with our secondary hypothesis (h2), the study did not find any difference in the participants’ sense of control in the two groups. After reviewing the literature (17), when the feeling of control relates to a choice, authors usually call it « autonomy » instead of « control ». Futures study could improve the precision of measurements by using authonomy-related validated scales (18).

There was no significant difference between the two groups regarding the desire to be exposed to a nudge again. This could be due to the rating scale in the questionnaire. A 5 or 7-point Likert scale could have shown significance.

All of these results lead to a broader reflection on the ethics of nudges and their potential danger. Nudging is considered by some as manipulation and by others as a legitimate solution to many social problems. For Thaler and Sunstein, nudge is about people’s well-being and should be transparent and publicly defensible. According to Beauvois, in order for nudge not to be considered as manipulation, the intentions of the authors and the functioning of nudge must be explicit and understood by the person exposed, thus being a transparent incentive.

Their significant effectiveness, the absence of a feeling of loss of autonomy in choice and the acceptability of nudge found significantly in the literature demonstrate the power of such an intervention.

### Strengths and limitations

This is a randomized, controlled study with a pre-registered protocol. The results are consistent with other interventional studies on the same topic (2,14,15) and give these results a strong external consistency in several contexts (application of different nudges, and to different populations, patients versus medical interns).

However, this is a study with a small number of participants. It would be interesting to extend this study to a larger number and to modify the questionnaire with numerical Likert scales instead of verbal binary scales. Participants reported that they were more likely to accept a proposed appointment following a call made from the practice’s telephone because they reported being more confident, which may constitute a bias. In addition, having met patients participating in the study beforehand as part of the investigator’s internship and having created a relationship of trust may have positively influenced adherence to the study and to vaccination during the telephone call.

## Conclusion

In this study, the nudge was significantly effective and highly accepted in both groups. Despite the significant importance of the boys who completed the vaccination as a result of the nudge, the participants did not express any loss of autonomy in choosing to be vaccinated. The study found that participants in the nudged group would like to be exposed to a nudge on similar topics such as vaccination again.

Furthermore, it would be interesting to correlate the effectiveness and acceptability of a nudge with a behaviour considered as normative. Normative behaviour is behaviour that conforms to norms (19),societal norms for example. Prevention posters make it possible to make behaviour normative (the population sees pro-HPV vaccination posters in the street, which makes this vaccination normative. However, they do not take the step to get vaccinated). The posters make the nudge normative and therefore acceptable to the target population (19). This could explain the failure of advertising campaigns and prevention posters if they are not correlated with a nudge.

The application of behavioural science has the potential to massively increase the prevalence of low-budget immunization nationwide. It is therefore important to continue to work in partnership with social psychology researchers. Research into the ethical nature of nudge must be a *prerequisite*, as participants do not realize that their behavior has been influenced. Indeed, they did not perceive any threat to their autonomy of choice, despite the nudge’s significant effectiveness. Decision-makers must therefore beware of the excesses of this type of intervention, which influences behavior despite the impression of complete autonomy.

## Data Availability

All data produced in the present study are available upon reasonable request to the authors

## Declaration

### Ethical approval

As an interventional study in the human and social sciences applied to the field of health, the study was classified out of scope of the Jardé law by the Ethics Committee (CPP) of Saint Etienne (ID RCB number: 2021-A01140-41 10.05.2021). A favorable opinion was issued by the Ethics Committee of the Non-interventional research (CERNI) of the Université Côte d’Azur (Number AVIS n° 2022-018). This work was the subject of a declaration of compliance with the reference methodology 004 to the CNIL n°2221990v0.

## Data availability

The data underlying this article will be shared upon request to the corresponding author.

## Referencies

1. Cancer du col de l’utérus : la couverture du dépistage et de la vaccination doivent progresser pour une meilleure prévention [Internet]. [cité 12 juill 2022]. Disponible sur: https://www.santepubliquefrance.fr/presse/2022/cancer-du-col-de-l-uterus-la-couverture-du-depistage-et-de-la-vaccination-doivent-progresser-pour-une-meilleure-prevention

2. Barbaroux A. Nudge. La place du sentiment de contrôle dans lefficacité et lacceptabilité des nudges en soins primaires [Internet]. Université Côte d’Azur; 2021 [cité 12 juill 2022]. Disponible sur: http://www.theses.fr/s312840

3. Adkisson R. Nudge: Improving Decisions About Health, Wealth and Happiness, R.H. Thaler, C.R. Sunstein. Yale University Press, New Haven (2008), 293 pp. The Social Science Journal. 1 déc 2008;45:700–1.

4. Sheeran P, Webb TL. The Intention–Behavior Gap. Social and Personality Psychology Compass. 2016;10(9):503–18.

5. Meszaros J, Ho CH, Corrales Compagnucci M. Nudging Consent and the New Opt-Out System to the Processing of Health Data in England. In 2019. p. 61–81.

6. Leclère C. Les nudges : un outil pour les politiques publiques ? Idées économiques et sociales. 2017;188(2):41–7.

7. Li M, Chapman GB. Nudge to Health: Harnessing Decision Research to Promote Health Behavior. Social and Personality Psychology Compass. 2013;7(3):187–98.

8. Korn L, Betsch C, Böhm R, Meier NW. Social nudging: The effect of social feedback interventions on vaccine uptake. Health Psychol. nov 2018;37(11):1045–54.

9. Brewer NT, Chapman GB, Rothman AJ, Leask J, Kempe A. Increasing Vaccination: Putting Psychological Science Into Action. Psychol Sci Public Interest. déc 2017;18(3):149–207.

10. Hagman W, Andersson D, Västfjäll D, Tinghög G. Public Views on Policies Involving Nudges. 2015;

11. Khodadadi AB, Hansen B, Kim YI, Scarinci IC. Latinx Immigrant Mothers’ Perceived Self-Efficacy and Intentions Regarding Human Papillomavirus Vaccination of Their Daughters. Womens Health Issues. juin 2022;32(3):293–300.

12. La vaccination contre les papillomavirus humains (HPV) étendue aux garçons [Internet]. [cité 18 juill 2022]. Disponible sur: https://www.ameli.fr/corse-du-sud/medecin/actualites/la-vaccination-contre-les-papillomavirus-humains-hpv-etendue-aux-garcons

13. Lovakov A, Agadullina E. Empirically derived guidelines for effect size interpretation in social psychology. European Journal of Social Psychology. 4 févr 2021;51:485–504.

14. Benoit L. Acceptabilité sociale et efficacité d’un nudge promouvant la vaccination antigrippale auprès de professionnels de santé. 2019;52.

15. Barbaroux A, Serati I. Effectiveness and acceptability of an opt-out nudge to promote influenza vaccination among medical residents in Nice, France: a randomized controlled trial [Internet]. medRxiv; 2022 [cité 12 sept 2022]. p. 2022.09.09.22279772. Disponible sur: https://www.medrxiv.org/content/10.1101/2022.09.09.22279772v1

16. Bonniot-Cabanac MC, Cabanac M, Fontanari JF, Perlovsky LI. Instrumentalizing Cognitive Dissonance Emotions. PSYCH. 2012;03(12):1018–26.

17. Skinner EA. A guide to constructs of control. Journal of Personality and Social Psychology. 71(3):549–70.

18. Wachner J, Adriaanse MA, De Ridder DTD. And How Would That Make You Feel? How People Expect Nudges to Influence Their Sense of Autonomy. Frontiers in Psychology [Internet]. 2020 [cité 15 août 2022];11. Disponible sur: https://www.frontiersin.org/articles/10.3389/fpsyg.2020.607894

19. Fehr E, Schurtenberger I. Normative foundations of human cooperation. Nat Hum Behav. juin 2018;2(7):458–68.

